# Liver Shape Analysis using Statistical Parametric Maps at Population Scale

**DOI:** 10.1101/2022.08.18.22278951

**Authors:** Marjola Thanaj, Nicolas Basty, Madeleine Cule, Elena P Sorokin, Brandon Whitcher, Jimmy D Bell, E Louise Thomas

## Abstract

**Background:** Morphometric image analysis enables the quantification of differences in the shape and size of organs between individuals.

**Methods:** Here we have applied morphometric methods to the study of the liver by constructing surface meshes from liver segmentations from abdominal MRI images in 33,434 participants in the UK Biobank. Based on these three dimensional mesh vertices, we evaluated local shape variations and modelled their association with anthropometric, phenotypic and clinical conditions, including liver disease and type-2 diabetes.

**Results:** We found that age, body mass index, hepatic fat and iron content, as well as, health traits were significantly associated with regional liver shape and size. Interaction models in groups with specific clinical conditions showed that the presence of type-2 diabetes accelerates age-related changes in the liver, while presence of liver fat further increased shape variations in both type-2 diabetes and liver disease.

**Conclusions:** The results suggest that this novel approach may greatly benefit studies aiming at better categorisation of pathologies associated with acute and chronic clinical conditions.

## Introduction

Despite improvements in global health [1], incidence of liver disease continues to rise, with deaths due to hepatic conditions increasing by 400% since the 1970s (British Liver Trust - https://britishlivertrust.org.uk/), making it the leading cause of death in those aged 35-49 years in the UK (ONS 2019 - https://www.ons.gov.uk/). Significant progress has been made in recent years in the use of non-invasive imaging methods to measure the pathological changes that are features of increasingly common liver conditions. This includes non-alcoholic fatty liver disease (NAFLD) [2, 3], fibro-inflammation [4, 5] and fibrosis [6]. The prevalence of these conditions, associated with obesity, insulin resistance and type-2 diabetes (T2D), are likely only to increase further given the current obesogenic environment. New approaches are needed to differentiate between those with mild disease, compared with those at risk of more significant conditions (cirrhosis/end stage liver disease), and particularly those who may experience accelerated disease processes [7]. One potential approach to address these issues is the implementation of novel morphometric methods to gain a deeper understanding of the processes underpinning the development and progression of many clinical conditions [8]. For instance, investigating whether changes beyond simple volume or fat measurements, such as liver shape, are associated with particular environmental risk factors, or whether they can be differentially related to the aetiology of a particular condition. These methods may potentially provide insight into different mechanisms of disease development and enable optimised treatment strategies to be developed.

Automated segmentation of the liver to produce image-derived phenotypes (IDPs) such as volume or fat deposition measurements are becoming more commonplace at scale as deep learning methods gain traction [9]. While these methods enhance our understanding of the liver at a population level, they are limited when it comes to providing additional knowledge regarding morphological, functional and regional variation in response to a particular condition.

Mapping organ segmentations to a standardised three-dimensional (3D) surface mesh, enables many thousands of measurements relating to variation in organ shape to be performed using statistical parametric maps (SPMs). A similar widely applied technique is statistical shape analysis, which transforms the 3D surface mesh measurements into a smaller number of principal components, known as shape parameters, has been used to characterise variations in organ shape across a population. These approaches have been successfully applied in neuroimaging [10, 11], abdominal computer tomography (CT) images [12, 13], and cardiac imaging [14, 15] and they have shown to be useful in identifying genetic interactions with cardiac pathology [16] and brain ageing [17]. However, they have been less frequently applied to abdominal organs, where morphological changes are known to take place in a variety of clinical conditions [18, 19].

In the current study we have applied SPM methods to determine morphological variations in the liver and their potential association with anthropometric traits and clinical conditions. We further investigated whether the emerging 3D liver mesh-derived phenotype can add value to the prediction of disease outcomes. Our study made three main contributions. The first contribution is that we investigate the impact of the population size and the robustness of the liver template construction. Specifically, we investigated how the template image and statistical parametric mapping are affected, providing valuable insights into determining the optimal number of subjects for the liver template to represent the broader cohort. We also examined the relevance of different participant samples in the template construction process. The second contribution and also a novelty of our work is that we extend the SPM method to the domain of liver image analysis. Here, we delve deeper into the application of SPM in liver image analysis and applied it to the UK Biobank dataset, which comprises a large-scale population-based cohort, resulting in increased statistical power. Through the linear regression model, we examined the impact of anthropometric, phenotypic and clinical conditions on regional geometry of the liver and visualised these findings on the template surface mesh. The third contribution is that we extracted shape features derived from the 3D mesh-derived phenotype by dimensionality reduction and evaluated whether these shape features were better predictors of disease outcomes than the conventional measurement of liver volume.

## Methods

### Data

The UK Biobank [20] is a population-based study in which 500,000 participants aged 40 to 70 years were recruited for deep phenotypic profiling. There is also a currently ongoing imaging sub-study, in which 100,000 of the participants have been recruited to undergo an imaging protocol including MRI of the brain, the heart, and the abdominal region. The abdominal scans include a neck-to-knee Dixon 3D acquisition that can be used to derive volumes of adipose tissue, skeletal muscle and abdominal organs. Full details regarding the UK Biobank abdominal acquisition protocol have previously been reported [21]. We processed and segmented the data using our automated methods [9]. In this study on liver morphology, we included 41,800 participants with Dixon MRI data acquired at the imaging visit, between 2014 and 2020 with data comprising imaging, health-related diagnoses and biological measurements.

Fully anonymized participant data was obtained through UK Biobank Access Application number 44584. The UK Biobank has approval from the North West Multi-Centre Research Ethics Committee (REC reference: 11/NW/0382) written informed consent was obtained from all participants prior to inclusion in the UK Biobank.

### Phenotype Definitions

Anthropometric measurements including age, body mass index (BMI), waist and hip circumferences were taken at the UK Biobank imaging visit and ethnicity was defined based on the continental genetic ancestry (https://pan.ukbb.broadinstitute.org). AST:ALT ratio, defined as the ratio of aspartate aminotransferase (AST) to alanine aminotransferase (ALT), commonly used to indicate presence of more advanced liver disease including fibrosis and cirrhosis [22, 23] was calculated from the biological samples taken at the initial assessment visit. The fibrosis-4 index (FIB-4), also designed to identify more advanced stages of liver disease and fibrosis in particular, was calculated as previously described [24] using age, AST, ALT and platelet count taken from the initial assessment visit. Diagnosis of liver disease and T2D was obtained from UK Biobank hospital records and self-reported information (see Disease Categories in supporting information). Due to the relatively limited number of scanned participants within the UKBB diagnosed with specific liver diseases, a broad umbrella definition of liver disease was implemented which included, alcoholic liver disease, fibrosis, cirrhosis, and chronic hepatitis.

### Quality Control

We included liver segmentations from an overall 41,800 participants. For details on the segmentation process and quality control refer to the supplementary data in [9]. Participants with missing clinical, anthropometric or biochemical data, as well as those with Dixon MRI datasets that did not have full anatomical coverage were excluded from the study, including organs with zero volume. More specifically, we removed 8,297 data that were missing ethnicity, BMI, WHR, AST, ALT, platelet count and liver IDPs. We also conducted quality control measures to determine potential extreme values in the liver volume and ensure the full anatomical coverage of the organs by visually examining values falling outside from randomly selected quantiles (0.1% and 99.9%) and excluding eight outliers. We visually inspected segmentations with 3D liver mesh-derived values to potentially identify extremely high values, resulting in the exclusion of 61 datasets with segmentation errors. Overall, from the initial 41,800 participants, 33,434 participants were included in the final analysis (20% of data excluded).

### Study Design

#### Template Definition

Deformation of an image to a standard organ template is a key part of MRI organ shape assessment. Given the potential variation in morphology, it is important to identify a suitable population sample size for constructing a template image [25]. To assess the impact of population size on template construction, we constructed three distinct templates using liver segmentations from a gender-balanced European ancestry cohort of 20, 100 and 200 participants with BMI<25 kg/m^2^ and low liver fat (<5%). The characteristics for each template population are provided in Supplementary Table S1. To test the 3 templates, we selected 500 participants, derived from the full cohort, with European genetic ancestry, aged between 46 and 62 years old, without any disease reported or diagnosed here [26] (Supplementary Table S2). We then registered the three liver templates to the 500-participant cohort and investigated the associations between the 3D mesh-derived phenotype and the anthropometric covariates across the three templates.

#### Association between mesh-derived phenotypes, IDPs and Disease

To assess the associations between the 3D mesh-derived phenotype, the anthropometric covariates and liver IDPs (volume, fat, iron), we first analysed the liver MRI data from the entire UK Biobank imaging cohort. The cohort of 33,434 participants was 97.6% European, 48.7% male and aged between 44 and 82 years old (Supplementary Table S3). To determine the potential association between disease and liver shape, we first selected diseases that are known from previous studies to impact liver health, and are associated with changes in liver fat accumulation or volume [9]. These included 449 participants with liver disease (207F/242M; 48-81 years old; BMI 18.6-43.8 kg/m²) and 1,780 participants with T2D (67% males; 46-82 years old; BMI 18.3-50.1 kg/m²) (Supplementary Table S4).

#### Prediction of disease outcomes

To determine whether the 3D mesh-derived phenotype was a better predictor of disease outcomes than the conventional measurement of liver volume, we identified 182 participants with liver disease (45% males; 45-78 years old; BMI 16.5-46.1 kg/m²) and 144 participants with T2D (61% males; 45-80 years old; BMI 19.9-47.9 kg/m²) that were diagnosed after the baseline imaging visit (see supporting information). We then identified a control cohort without any reported conditions and designed a case-control study for each disease population, achieving a 364 case-control cohort with liver disease and 288 case-control cohort with T2D. The control cohort was chosen by matching one individual with every case by age (± 1 year), gender and BMI (± 2 kg/m²) using the R package *ccoptimalmatch* [27].

#### Image Registration and Mesh Construction

The process for template construction of the liver has been previously described [28]. Here, we constructed three distinct templates using liver segmentations from 20, 100 and 200 subject-specific volumes in order to evaluate the impact of cohort size on template construction. It also allows us to test if cohort size influenced the statistical associations in our mesh-based analysis. We constructed surface meshes from each template using the marching cubes algorithm and smoothed using a Laplacian filter [29]. The template construction was performed using ANTs software (https://picsl.upenn.edu/software/ants) with mutual information as the similarity metric and the B-spline non-rigid transformation. Briefly the process of the template construction is performed in two stages: affine registration to account for translation, rotation, scaling and shearing, and non-rigid registration to account for local deformation using the symmetric image normalisation (SyN) method with mutual information as the similarity metric [30, 31]. The analysis was performed using “antsMultivariateTemplateConstruction2.sh” script provided from ANTs, with the following default parameters: -i (iteration limit) = 4, -g (gradient step size) = 0.25, -k (number of modalities) = 1, -w (modality weight) = 1. The rest parameters were customised depending on the machine used, image dimension and the metrics applied, including: -d (image dimension) = 3, -j (number of CPU cores) = 10, -c (control for parallel computation) = 2, -q (max iteration for each pairwise registration) = 100×70×50×10, -n (NBiasFieldCorrection of moving image) = 0, -r (do rigid body registration of inputs to the initial template) = 1, -m (similarity metric) = MI and -t (transformation model) = BSplineSyN.

Surface meshes were first constructed from each subject’s segmentations using marching cubes algorithm and smoothed using a Laplacian filter. Then the template-to-subject registration was performed by first applying rigid registration to remove the position and orientation difference between all subject-specific surfaces and template surfaces and an affine transformation with nearest neighbour interpolation was computed between template and subject segmentations. The resulting affine transformations were used to warp the template to the subject’s space. The template segmentation is then mapped into each subject segmentation by computing a non-rigid transformation modelled by a free-form deformation, based on B-Splines, with label consistency as the similarity metric between the subject and template liver segmentations [32]. To enable subject comparison with vertex-to-vertex correspondence, the template mesh is then warped to each subject mesh using the deformation fields obtained from the non-rigid registration. Hence, all surface meshes are parameterised with the same number of vertices (approximately 18,000). This ensures that each vertex maintains approximate anatomical accuracy and consistency across all subjects, while preserving the size and shape information for subsequent analyses [29].

To determine the regional outward or inward adaptations in the liver surface in comparison to an average liver shape, the surface-to-surface (S2S) distance, a 3D mesh-derived phenotype for each subject was measured. This was achieved by computing the signed distance between each vertex in the template mesh and each corresponding vertex in the subjects’ mesh. This indicates positive distances for outward expansion in the subject’s vertices compared to template vertices and negative distances for inward shrinkage in the subject’s vertices. All the steps for the template-to-subject registration were performed using the Image Registration Toolkit (IRTK) (https://biomedia.doc.ic.ac.uk/software/irtk). After conducting the described manual quality control process, which involved identifying extremely high S2S values, we found that all the values fell within the range of −48.3 to 70.5 mm. This is to ensure that the organ sizes were within an expected range and to suggest that there were no significant segmentation errors, such as the inclusion of surrounding tissues in the liver segmentations.

#### Mass Univariate Regression

Associations between the S2S values and anthropometric variables were modelled using a linear regression framework. To enhance the detection of spatially contiguous signals and discriminate them from noise, we utilised threshold-free cluster enhancement (TFCE) [33]. TFCE not only provides improved sensitivity and stability compared to other cluster-based techniques but also identifies local maxima in the resulting significance map that is not possible in other enhancement and thresholding techniques [14, 33]. A permutation testing was then performed on the TFCE maps and the derived TFCE p-values were corrected to control the false discovery rate (FDR), as previously described [28]. Specifically, we performed mass univariate regression (MUR) analysis using the R package *mutools3D* [34] and adjusted for multiple comparisons by applying the FDR procedure [35] to all the TFCE p-values derived from each vertex using 1,000 permutations. The estimated regression coefficients 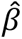
 for each of the relevant covariates and their related TFCE-derived p-values were then displayed at each vertex in the mesh on the whole 3D liver anatomy, providing the spatially-distributed associations. Regions of the liver exhibiting significant associations (p-values < 0.05) between variables were identified, and the estimated regression coefficients 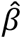
 for each relevant covariate within those regions were reported. The MUR model for deriving associations between clinical parameters and a 3D phenotype is outlined in Supplementary Fig. S1.

To determine which factors influence the design and performance of the liver template, we used a regression model to address: (1) how many participants are required to construct a representative liver template, (2) whether the template population size affected the associations between the S2S and the anthropometric covariates, (3) which factors have an impact on regional S2S distances and (4) how are the changes in S2S distances linked to liver disease and T2D.

We constructed three models adjusting for additional covariates. **Model 1** was adjusted for age, gender, ethnicity, body mass index (BMI) and waist-to-hip ratio (WHR), liver fat (referred to as proton density fat fraction (PDFF)) and liver iron concentration with correction to control the FDR. To investigate the morphological changes related to liver function **Model 2** had all the covariates from model 1 plus AST:ALT, FIB-4 index and disease conditions. We further adjusted with interaction terms between age and disease status and between liver fat and disease. In order to test whether there is a circadian effect in the liver morphology, **Model 3** included all the covariates from model 2 plus time of the day for the MRI scan, discretised into hours of the day.

#### Predictive Model

To determine whether S2S distance improves the prediction of disease outcomes prospectively, we used a logistic regression model. This model allowed us to investigate the associations between liver volume as well as the S2S values from the baseline imaging visit and the occurrence of disease outcomes in two distinct case-control cohorts: one comprising individuals with liver disease and the other with T2D.

Due to having a large number of S2S values for small population groups, we first calculated the sparse principal component analysis (SPCA) using the R package *sparsepca* [36] and extracted principal component scores representing the shape features of the S2S distances for each disease case-control group that were diagnosed after the baseline imaging visit. We utilised the principal component scores for each individual corresponding to the modes that summarised 90% of the cumulative variation for each group. We then performed this analysis in two models. In the first model (the volume model), the disease outcome was regressed on age, gender, ethnicity, BMI, WHR, AST/ALT, FIB-4 index, liver volume, PDFF and iron concentration. In the second model (the S2S model), we included all the covariates from the volume model, adding the principal component scores of the S2S distances for each disease group.

Predictive modelling was performed using the R package *caret* [37]. Model training was conducted with leave-one-out cross validation for each group. Our model performance was evaluated using several metrics, including the Area Under the Curve (AUC) of the Receiver Operating Characteristic (ROC) curve, the F1 score, accuracy, and sensitivity/specificity. Additionally, we employed Delong’s test to compare the AUC of the ROC curves from S2S and liver volume models [38].

## Results

### Template Consistency

We constructed three separate template meshes using gender-balanced cohorts of 20, 100 and 200 participants and computed the distances between each template mesh for each subpopulation (Supplementary Fig. S2). The results showed that cohort size had little impact on the shape of the template, with differences less than 8mm, especially for the templates constructed using 100 participants compared with the 200-participant template. More specifically, the median absolute distance between the 20-participant and 200-participant templates was found to be −1.1 (IQR: 3.2) mm, whereas the median distance between the 100-subject and 200-subject templates was even smaller (−0.4 (1.8) mm). To further examine the relevance of different participant samples in the template construction process, we constructed five templates, each constructed from different samples drawn from a population of 20 participants each. The Dice coefficients of the template images for the 20-participant template experiment consistently demonstrates a high level of overlap across the distinct cohorts (Supplementary Table S5). It is important to note that when constructing templates using larger cohort sizes (e.g., 100 or 200 participants), it is expected that the variability will be reduced due to the averaging effect. Based on these findings, we are confident in the robustness and consistency of our template construction process.

We further investigated for each template the associations between S2S distances and anthropometric variables, adjusting for the covariates in Model 1 to examine how the statistical parametric mapping is influenced across the three templates. We only looked at the associations between BMI and WHR with S2S distances, as only these variables exhibited statistically significant associations. Here we visually presented the 3D SPMs, with the TFCE corrected p-values, of BMI and WHR with the S2S distance on the 500-participant cohort (Supplementary Fig. S3) and presented the significance areas of their associations across the three templates (Table 1). By combining qualitative and quantitative assessments, we showed that the distribution of the corrected p-values were consistent across all three different templates and that there was no apparent difference in the areas of association between BMI and WHR with S2S distances across the three templates.

**Table 1.** Significance areas from the association between BMI and WHR with S2S distances on a 500-participants cohort, in the MUR model using a template with 20, 100 and 200 participants. The significance area is the percentage of vertices on the liver mesh where the regression coefficients are statistically significant (p < 0.05) after adjustment for multiple comparisons. The total area has been split into areas of negative (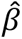
 < 0) and positive (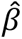
 > 0) associations.

| Significance area | 20-participant template |  | 100-participant template |  | 200-participant template |  |
| --- | --- | --- | --- | --- | --- | --- |
|  | BMI | WHR | BMI | WHR | BMI | WHR |
| Total | 58.08% | 14.48% | 55.20% | 18.28% | 56.73% | 12.12% |
| $\hat{\beta} < 0$ | 2.74% | 4.79% | 3.28% | 7.42% | 2.66% | 4.67% |
| $\hat{\beta} > 0$ | 55.34% | 9.69% | 51.92% | 10.85% | 54.07% | 7.46% |

To test template consistency on a disease population, all three templates were registered on a cohort of 449 participants with liver disease and the 3D S2S phenotype computed between template and participants’ surface. We then modelled the associations between the S2S distances and anthropometric variables adjusting for the covariates in model 1. The TFCE corrected p-value maps on the cohort with liver disease were consistent across the three templates, with little difference in the significance area for the association between BMI and S2S distances (97.58% using the 20-participant template, 97.46% using the 100-participant template and 96.43% using the 200-participant template) (Supplementary Fig. S4 and Table 2).

**Table 2.** Significance areas from the association between BMI and WHR with S2S distances on a cohort with liver disease (N=449), in the MUR model using a template with 20, 100 and 200 participants. The significance area is the percentage of vertices on the liver mesh where the regression coefficients are statistically significant (p < 0.05) after adjustment for multiple comparisons. The total area has been split into areas of negative (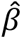
 < 0) and positive (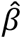
 > 0) associations.

| Significance area | 20-participant template |  | 100-participant template |  | 200-participant template |  |
| --- | --- | --- | --- | --- | --- | --- |
|  | BMI | WHR | BMI | WHR | BMI | WHR |
| Total | 97.58% | 91.02% | 97.46% | 90.31% | 96.43% | 90.98% |
| $\hat{\beta} < 0$ | 0.01% | 0.01% | 0% | 0% | 0% | 0% |
| $\hat{\beta} > 0$ | 97.57% | 91.01% | 97.46% | 90.31% | 96.43% | 90.98% |

### Associations with Anthropometric Characteristics, Liver IDPs and Disease

As the liver template was relatively insensitive to the number of participants included, we performed all subsequent analyses using the 200-participant template. We proceeded to register the template on the full cohort (N=33,434), computing S2S distances between the template and surface of each individual liver mesh and performed MUR analysis adjusting for the covariates in Model 2.

A summary of the model for the whole cohort, representing the regression coefficients and the significance areas on the liver, is provided in Table 3 and Supplementary Fig. S5. The SPMs that represent associations between S2S distances and the anthropometric measurements and liver IDPs with units in standard deviations for each covariate, are shown in Fig. 1.

**Figure 1.**
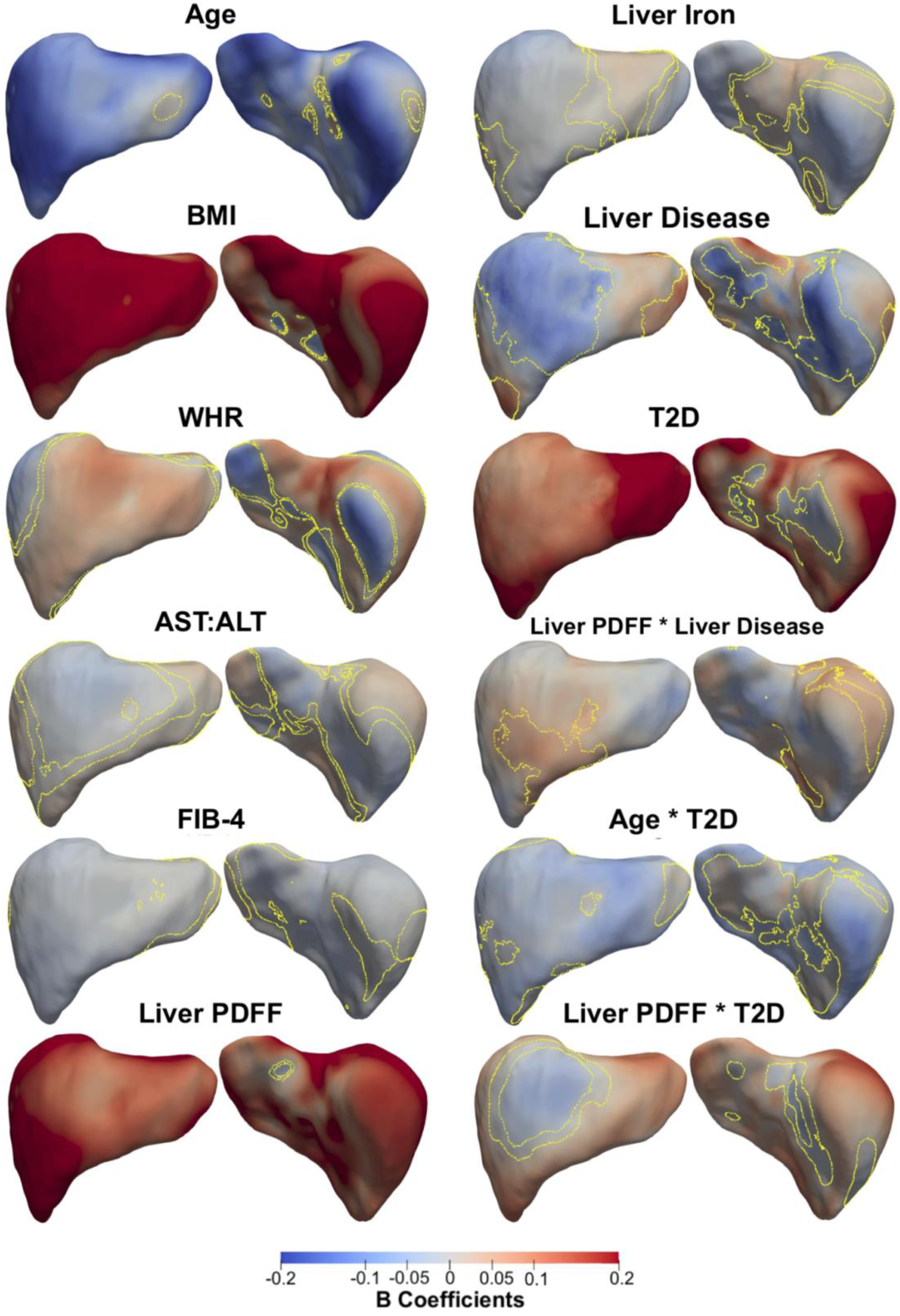
Three-dimensional statistical parametric maps (SPMs) of liver morphology, two projections are shown for each SPM providing anterior (left) and posterior (right) views of the liver. The SPMs show the local strength of association for each covariate in model 2 with S2S distances on the full cohort (N=33,434). Yellow contour lines indicate the boundary between statistically significant regions (p < 0.05) after correction for multiple testing, with positive associations in red and negative associations in blue. Standardised regression coefficients are shown with units in standard deviations for each covariate. BMI: body mass index, WHR: waist-to-hip ratio, AST:ALT: aspartate aminotransferase/alanine aminotransferase ratio, FIB-4: Fibrosis-4 score, Liver PDFF: Liver percentage density fat fraction, T2D: type-2 diabetes.

**Table 3.** Significance areas for covariates in the MUR model between the anthropometric covariates and liver IDPs (N=33,434) in model 2. The total area has been split into areas of positive and negative associations. The regression coefficients are presented as median (interquartile range - IQR) and the significance areas as a percentage (%) of the vertices. Where BMI: body mass index, WHR: waist-to-hip ratio, AST:ALT: aspartate aminotransferase/alanine aminotransferase ratio, FIB-4: Fibrosis-4 score, Liver PDFF: Liver percentage density fat fraction, T2D: type-2 diabetes, ns: not significant.

| | $\hat{\beta} < 0$ | | $\hat{\beta} > 0$ | | Total |
| --- | --- | --- | --- | --- | --- |
|  | Beta coefficients | Significance area | Beta coefficients | Significance area | Significance area |
| **Age (yrs.)** | -0.11 (0.06) | 96.63% | 0.02 (0.04) | 1.46% | 98.10% |
| **BMI ($kg/m^2$)** | -0.08 (0.07) | 1.61% | 0.30 (0.22) | 97.82% | 99.43% |
| **WHR** | -3.88 (4.02) | 33.99% | 3.87 (3.65) | 58.11% | 92.10% |
| **AST:ALT** | -0.32 (0.32) | 35.17% | 0.30 (0.29) | 48.05% | 83.22% |
| **FIB-4** | -0.22 (0.17) | 82.62% | 0.23 (0.13) | 2.09% | 84.70% |
| **Liver PDFF (%)** | -0.03 (0.02) | 0.17% | 0.26 (0.10) | 99.65% | 99.82% |
| **Liver Iron ($mg/g$)** | -0.59 (0.74) | 58.00% | 0.34 (0.32) | 24.99% | 82.98% |
| **Liver disease** | -2.13 (2.95) | 21.90% | 1.95 (2.43) | 25.14% | 47.05% |
| **T2D** | -0.61 (0.77) | 5.35% | 2.42 (1.94) | 86.40% | 91.76% |
| **Age \* Liver** | ns | ns | ns | ns | ns |

**Table 3.** Significance areas for covariates in the MUR model between the anthropometric covariates and liver IDPs (N=33,434) in model 2.
| disease |  |  |  |  |  |
| --- | --- | --- | --- | --- | --- |
| **Liver PDFF \***<br>**Liver disease** | -0.09 (0.01) | 0.09% | 0.09 (0.03) | 12.59% | 12.68% |
| **Age \* T2D** | -0.03 (0.02) | 71.23% | 0 | 0% | 71.23% |
| **Liver PDFF \***<br>**T2D** | -0.06 (0.04) | 6.24% | 0.10 (0.08) | 82.84% | 89.08% |

Lower S2S distances were associated with greater age over 96.63% of the liver, with a median change of −0.11 mm/year, while BMI and WHR had statistically significant positive associations with S2S distances, covering 97.82% and 58.11% of the liver, respectively. The AST:ALT ratio showed mostly statistically significant positive association with S2S distances in the anterior part of the left lobe and the posterior part of the right lobe, with a median difference of 0.30 mm (significance area = 48.05%). FIB-4 index on the other hand showed a median S2S distance of −0.22 mm (significance area = 82.62%). Liver PDFF was positively associated with S2S distances, showing median outward shape variations of 0.26 mm/%, whereas liver iron concentration was associated with S2S distances of −0.59 mm/(mg/g) in the anterior part of the right lobe and the posterior part of the left lobe and a median 0.34 mm/(mg/g) in the anterior part of the left and caudate lobe. Additionally, we included MRI scan time as an additional covariate in the model since liver size is known to vary during the day [9], but this had no apparent effect on any of the associations (Supplementary Table S6, Supplementary Fig. S6).

A diagnosis of liver disease was associated with a median S2S of −2.13 mm when compared to the controls (significance area = 21.90%) in the anterior part of the right lobe as well as at the posterior part of left and right lobe and a median of 1.95 mm (significance area = 25.14%) in the anterior part of the left lobe. T2D was positively associated with S2S distances, with a median of 2.42 mm for participants with T2D covering a significance area of 86.40% of the liver. The time of day at which the MRI scan was conducted had no effect on the associations between S2S and T2D, although we observed a reduction in the significance area for the associations between S2S and liver disease (significance area = 28.34%, Supplementary Table S6, Supplementary Fig. S6).

We undertook further analysis to determine whether there was an interaction between clinicalstate and factors such as age and liver PDFF adjusted for all covariates in Model 2. Our results varied according to the disease of interest. While there were no significant associations for the interaction between age and liver clinical condition, we found a median association of −0.14 mm/year in T2D participants, compared with −0.11mm/year in non-T2D participants, over a similar anatomical region. The interaction term between age and T2D in this model was significantly different from zero, with a significance area = 71.23% (Table 3 and Fig. 1). The association between age and S2S distances in participants with and without T2D are directly compared in Fig. 2, where participants diagnosed with T2D display accelerated decreases in the anterior part of the left and right lobe as well as at the posterior part of left and right lobe of the liver.

**Figure 2.**
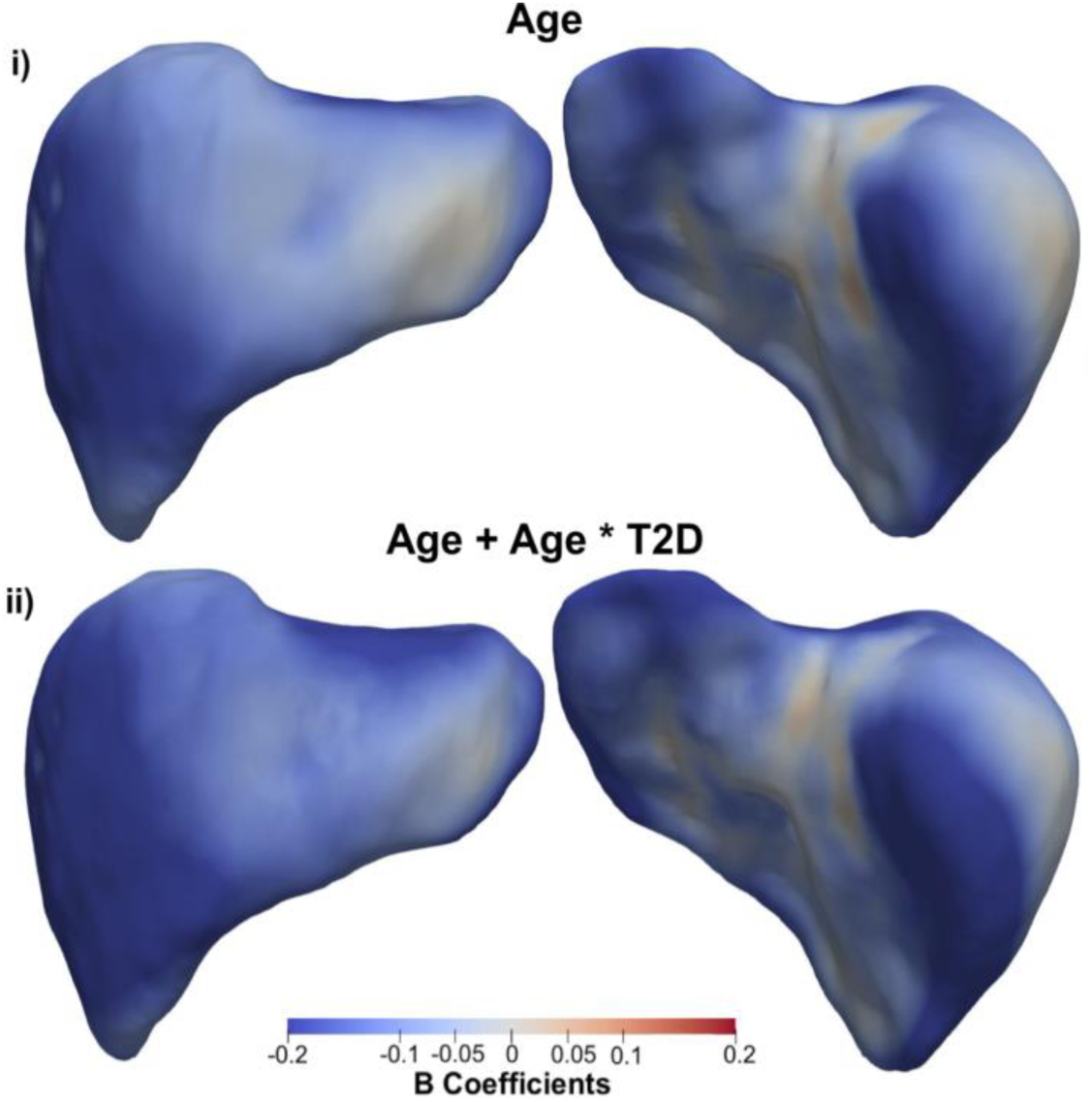
Three-dimensional statistical parametric maps (SPMs) of liver morphology, projections are anterior (left) and posterior (right). The SPMs show the local rate of change as a function of age for S2S distances in participants (i) without T2D versus those (ii) with T2D on the full cohort (N=33,434). Positive associations are in red and negative associations in blue. Standardised regression coefficients are shown with units in standard deviations.

The presence of liver PDFF in participants with liver disease resulted in an additional median variation of 0.09 mm/% over an area 12.59% of the liver, in addition to the median variation of 0.26 mm/% associated with the main effect of liver PDFF (Table 3 and Fig. 1). Interestingly this effect was no longer significant after including scan-time as an additional covariate in the model (Supplementary Table S6, Supplementary Fig. S6). A change of similar magnitude, over a much larger proportion of the liver was observed for the interaction between liver PDFF and T2D (Table 3 and Fig. 1). Here we observed an accelerated increase in S2S distances with a median change of 0.10 mm/%, over the majority of the liver surface area (significance area = 82.84%), in addition to the median increase of 0.26 mm/% for the main effect of liver PDFF. The rates of change in S2S distances due to changes in liver PDFF for participants with liver disease only, with T2D only and those without either disease are directly compared in Fig. 3. The local variations associated with liver PDFF fluctuates significantly with disease diagnosis. Participants diagnosed with liver disease (Fig. 3ii) display accelerated increases in S2S distances in the anterior and posterior parts of the right lobe with increasing liver PDFF, with slight decreases in the rate of change in both the anterior and posterior left lobe when compared to participants without either liver disease or T2D. Participants with T2D (Fig. 3iii) display accelerated increases in S2S distances in the anterior and posterior right lobe and the posterior left lobe when compared to participants without T2D, and display substantial decreases in the rate of change in S2S distances in the anterior left lobe when compared to participants who have been diagnosed with liver disease but not T2D or participants who have not been diagnosed with either liver disease or T2D.

**Figure 3.**
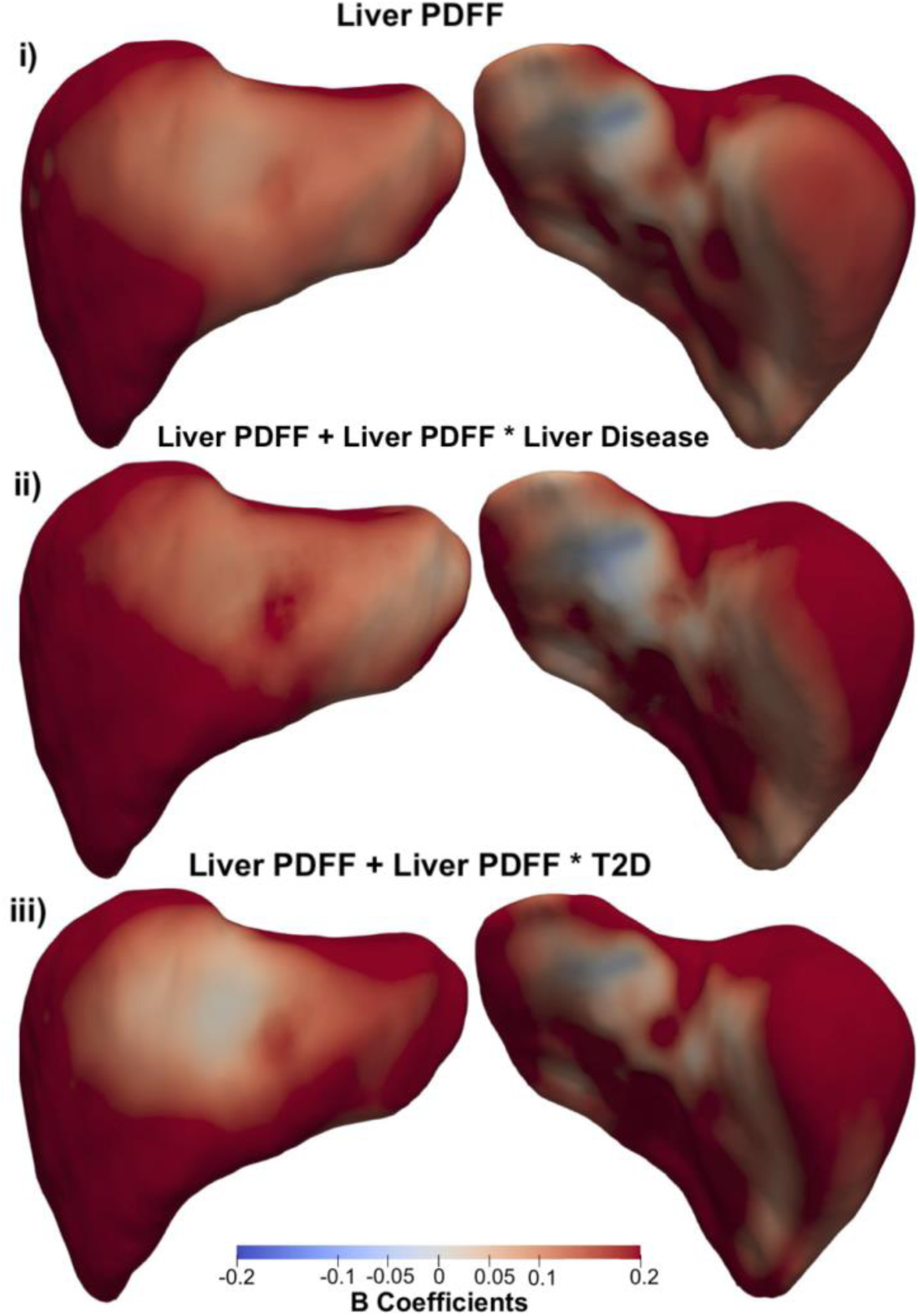
Three-dimensional statistical parametric maps (SPMs) of liver morphology, projections are anterior (left) and posterior (right). The SPMs show the rate of change as a function of liver PDFF for S2S distances in participants (i) without liver disease or T2D, (ii) with liver disease only and (iii) with T2D only on the full cohort (N=33,434). Positive associations are in red and negative associations in blue. Standardised regression coefficients are shown with units in standard deviations.

### Predictive Analysis

We investigated whether S2S distances add value to disease prediction beyond those obtained using liver volume. We compared the performance of two models; one including age, gender, ethnicity, BMI, WHR, AST:ALT, FIB-4 index, liver PDFF, liver iron and liver volume (the volume model); the other including age, gender, ethnicity, BMI, WHR, AST:ALT, FIB-4 index, liver PDFF, liver iron, liver volume and the principal component scores of the S2S distances (the S2S model), for the liver disease (N=364) and T2D (N=288) case-control cohorts. We found that the liver volume model achieved an AUC=0.57 and accuracy=0.54 (sensitivity/specificity=0.42/0.66) for liver disease prediction and AUC=0.64 and accuracy=0.62 (sensitivity/specificity=0.54/0.70) for T2D prediction (Table 4). The first 40 modes of the SPCA were sufficient to describe over 90% of the S2S distances in both cohorts, thus the first 40 scores in each cohort were used as independent variables in the model. The S2S model improved the prediction of liver disease achieving an AUC of 0.61, accuracy of 0.59 and sensitivity/specificity values of 0.57/0.60. However, when comparing the S2S and the volume models, the improvement was not statistically significant (p=0.1). Additionally, there was no statistically significant improvement in T2D (AUC=0.64, accuracy=0.62, sensitivity/specificity=0.59/0.64) compared to the model with liver volume.

**Table 4.** Predictive models trained with leave-one-out cross validation for both liver disease (N=364) and T2D (N=288) case-control groups. Each cell contains the area under the curve (AUC) with 95% confidence intervals (CI) in parentheses, F1 score and accuracy with sensitivity and specificity in parentheses.

| Case -<br>Control<br>Cohort | Models |  |  |  |  |  |
| --- | --- | --- | --- | --- | --- | --- |
|  | Volume |  |  | S2S |  |  |
|  | AUC (95% CI) | F1<br>score | Accuracy<br>(Sensitivity /<br>Specificity) | AUC (95% CI) | F1<br>score | Accuracy<br>(Sensitivity /<br>Specificity) |
| **Liver**<br>**disease** | 0.57 (0.50-0.62) | (0.59) | 0.54<br>(0.42 / 0.66) | 0.61 (0.55-0.67) | (0.60) | 0.59<br>(0.57 / 0.60) |
| **T2D** | 0.64 (0.58-0.71) | (0.65) | 0.62<br>(0.54 / 0.70) | 0.64 (0.57-0.70) | (0.62) | 0.62<br>(0.59 / 0.64) |

Supplementary Fig. S7 shows the increase in AUC with the increasing numbers of modes, from 1 until 40 for the prediction of liver disease and T2D. Notably, the S2S model for liver disease prediction reached its peak performance when utilising 21 modes, resulting in an AUC of 0.63 (95% confidence interval (CI): 0.57-0.69) and an F1 score of 0.64. This improvement was statistically significant (p=0.013), with an accuracy of 0.63 and sensitivity/specificity values of 0.60/0.65. Furthermore, we observed a slight enhancement in T2D prediction using the S2S model with 11 modes, resulting in an AUC of 0.67 (with 95% CI of 0.60 to 0.73) and an F1 score of 0.63. However, this improvement was not statistically significant (Supplementary Fig. S7).

## Discussion

In this study, we mapped local shape variations across the liver and determined how these changes were associated with anthropometric, phenotypic and health traits. To achieve this we constructed surface meshes from liver segmentations of 33,434 participants from the UK Biobank. Previous studies using similar SPMs have suggested that this is a useful technique in neuroimaging [10] and cardiac imaging [14], enabling the associations between phenotypic and genetic variation in specific anatomical regions to be mapped [16].

We constructed a representative liver template, and showed that a 200-participant template was sufficient to represent the broader cohort. Indeed, the number of participants included in the template construction did not impact the power of the statistical analysis across a 500-participant test cohort, or a second cohort of 479 participants with liver disease. This is in line with previous studies that found a cohort with 100 participants was sufficient to construct a representative cardiac template to investigate the shape of the left ventricle [29].

Liver size has been explored extensively using a variety of approaches from autopsy measurements [39], CT [40], ultrasound [41], and MRI [19], as well as regression-based algorithms designed to predict liver size based on body surface area [42]. Given accurate assessment of liver volume is essential for many aspects of hepatic surgery and determining disease progression [43], suitable methods are needed. However, until recently, the manual annotation required to make true volumetric measurements of the liver from CT and MRI images has been extremely time consuming. Imaging studies tended to rely on more easily measured metrics, such as liver span or diameter [44, 45], or calculation of volume indices from the measurement of multiple diameters [46]. Consequently, these approaches limit in depth morphometric assessment and only provide information associated with overall changes to liver size or volume. The SPM method implemented in the current study demonstrates significant regional variations in liver shape associated with anthropometric variables and disease status, including simultaneous inwards and outwards adaptations. These novel phenotypic variables may be useful in longitudinal population studies, as well as determining trajectories of progression in aggressive clinical conditions, including monitoring liver cirrhosis and hepatic oncology.

While studies of liver volume have generally focussed on patient populations, there is increasing interest in understanding how hepatic volume and form is influenced by age, anthropometry and metabolic markers in the wider population [9, 46]. Despite this, few studies employ methods that enable precise measurements of these parameters, particularly with regard to regional variation in liver shape and size. In the present study we observed that decline in the liver S2S distances were associated with increasing age. This is in agreement with previous observations, by ourselves and others, that overall liver volume decreases with age [9, 41, 47]. However, there are some ultrasound reports suggesting liver size increases with age [44]. This discrepancy may relate to variations in methodology since ultrasound measurements of liver diameter may not reflect overall changes in liver volume. This clearly reinforces the importance of absolute volumetric measurements, which, when combined with statistical parametric mapping, enables simultaneous extraction of global and local changes.

Additionally, we found a strong and distinct regionality in liver morphometry which was associated with body composition and liver PDFF. Specifically, we found that higher BMI and WHR were strongly associated with positive S2S distance, in line with others who have reported a positive correlation between liver size and anthropometric variables [45, 46]. We also found that higher liver PDFF was significantly associated with positive S2S distances, suggesting that hepatic fat is associated with both liver size and shape, with some clear regional variations. We further explored whether the time of day the participants were scanned was associated with S2S distances, given we have previously shown this to be associated with fluctuations in liver volume [9]. However, we did not find a measurable effect.

We investigated whether conditions with known involvement of hepatic function had discernible effects on our S2S measurements. For this we selected T2D, commonly associated with increased deposition of liver PDFF, and subjects with known liver conditions, which we expected to be associated with a more adverse phenotype. We found that T2D was associated with outward shape variations in the liver after adjusting for PDFF, suggesting that T2D affects liver morphology. It is well recognised that T2D is associated with a range of liver conditions, with the prevalence of NAFLD in patients with T2D reported to be 55% and NASH 37.3% [48], substantially higher than the proportion of individuals in the general population with NAFLD (19.9%) [3] or NASH (2.2%) [49]. Given the clinical heterogeneity of our current T2D cohort, in terms of time of diagnosis and medication, as well as the possibility of collider bias or reverse confounding, it is impossible to identify causal mechanisms for the observed variation in S2S distances. Interestingly when we considered the interaction between age and disease, we found no statistically significant interaction for liver disease, but there was a significant interaction between age and the presence of T2D. We also considered whether the interaction between disease and the presence of liver PDFF was associated with S2S distances. Moreover, the variations covered a larger proportion of the liver in T2D compared with liver disease. This may suggest that the hepatic tissue in T2D retains its overall relative plasticity (i.e. less fibrotic-cirrhotic tissue), while in liver disease there may be regions that have reduced capacity to accumulate fat or lost their plasticity and thus be less responsive to geometrical changes. Further work is needed to determine how these changes may be utilised to improve diagnosis or monitoring of disease progression. Future work in patients with biopsy-characterised hepatic tissue should help to shed light on the heterogeneity of response to the interaction between liver fat accumulation and liver health status.

We further identified regional variations in liver morphometry that are associated with liver disease. Specifically, we observed an inward shape variation at the anterior part of the right lobe, and posterior parts of the left and right lobes accompanied by an outward increase in liver S2S distances in the anterior part of the left lobe in participants diagnosed with liver disease. Previous studies have suggested that statistical shape modelling is a viable approach for enhancing the understanding of the liver shape variations linked to the stage fibrosis and even predicting it [13, 50]. With limited outcome and longitudinal data in the current study, the clinical significance of these changes, particularly the simultaneous regional inward and outward deformations in S2S distances are unclear. However, histological and radiological studies of the liver in patients with cirrhosis have shown that the degree of volume reduction and fibrosis is greater in the right lobe compared to the caudate lobe (which reportedly expands) [51]. This suggests regional changes in S2S distances may reflect physiological processes in the liver. It is well established that many diseases do not progress uniformly across the liver, with differences reported within different zones (periportal, mid-lobular and pericentral) of the liver lobule, which may reflect populations, different cell types, metabolic function and differences in blood flow [52]. Whilst it is premature to adjudicate a mechanism responsible for the changes described in the current study, the regional shape differences associated to both AST:ALT and FIB-4, hinting at hepatocellular changes underpinning the variation in S2S distances.

We assessed the predictive performance using shape features derived from the S2S distances on the case-control cohorts with liver disease and T2D. We aim to determine whether these shape features can add to prediction of disease beyond those obtained using conventional volumetric measurements. We demonstrated that the model using the shape features of the S2S distances improved the prediction of liver disease, however, there was no improvement in T2D compared to the model with liver volume. Our methods using the shape features, particularly in which histology is available, may provide additional information to confirm the utility of our approach in monitoring disease and potentially predicting outcomes. This in turn would open up the possibility of applying this methodology, in conjunction with other techniques to determine and predict the overall trajectory of progression of disease and identify those subjects requiring closer monitoring and more aggressive forms of treatment. Future work is also needed to explore variations in liver morphometry by condensing the entire coordinate matrix or deformation matrix into most distinct principal component modes to categorise population variations, which could be used in genetic association studies to enhance our understanding of chronic liver disease [17, 53].

Our study was not without limitations. To ensure sufficient numbers of participants in the liver disease group, we included all participants in the imaging cohort who had a diagnosis of liver disease, regardless of aetiology (alcoholic, toxic and inflammatory liver disease, hepatitis, fibrosis and cirrhosis). This precludes us from a more in-depth granular analysis, although our data does suggest that hepatocellular damage, particularly in more advanced disease stages, resulted in significant S2S changes across the liver. Variation in disease aetiology, the point of disease progression and the impact of on-going treatment may further confound the interpretation of our observations in the liver disease cohort. Furthermore, this study has only 3,088 follow-up data since the imaging visit, which limits the identification of more severe cases and may limit the predictive power. Additional longitudinal measurements will need to be required to assess age-related changes in disease cohorts.

## Conclusion

This study demonstrates that methods to assess changes in liver morphology, beyond simplistic volumetric analysis, can be applied at scale. In a population-based study we show that inter- and intra-subjects’ morphometric variations are associated with age, body composition and liver phenotypes, as well as disease. Moreover, morphometric scores were shown to improve the prediction of liver disease over-and-above conventional measures of liver volume. The approach developed here will allow large-scale studies of patient-based cohorts, enable disease-specific changes in morphology to be defined and tracked during both progression and remission and facilitate disease prediction and stratification.

## Data Availability

Researchers may apply to use the UKBB data resource by submitting a health-related research proposal (https://www.ukbiobank.ac.uk).

## Declarations

### Competing interests

M.C. and E.P.S. are employees of Calico Life Sciences LLC. M.T., N.B., B.W., J.D.B. and E.L.T. declare no competing interests.

### Ethics approval and consent to participate

The data resources used in this study have approval from ethics committees. Full anonymised images and participants metadata from the UK Biobank cohort was obtained through UK Biobank Access Application number 44584. The UK Biobank has approval from the North West Multi-Centre Research Ethics Committee (REC reference: 11/NW/0382), and obtained written informed consent from all participants prior to the study. All methods were performed in accordance with the relevant guidelines and regulations as presented by the relevant authorities, including the Declaration of Helsinki https://www.ukbiobank.ac.uk/learn-more-about-uk-biobank/about-us/ethics.

### Consent for publication

Not applicable.

### Availability of data and materials

The data that support the findings of this study are available from the UK Biobank (https://www.ukbiobank.ac.uk), but restrictions apply to the availability of these data, which were used under license for the current study, and so are not publicly available. Data are however returned by us to the UK Biobank where they will be fully available on request.

### Funding

This study was funded by Calico Life Sciences LLC, who provided the financial means to allow authors to carry out the study. The finding bodies played no role in the design of the study and collection, analysis, and interpretation of the data and the writing of the manuscript.

### Authors’ contributions

J.D.B., E.L.T., M.T. and M.C. conceived the study. J.D.B., B.W., E.L.T., N.B. and M.T. designed the study. M.T., N.B., B.W., E.P.S. and M.C. implemented the methods and performed the data analysis. M.T. defined the disease and physiological condition categories. M.T. performed the image and statistical analysis. E.L.T., B.W., M.T., J.D.B., and N.B. drafted the manuscript. All authors read and approved the manuscript.

## Acknowledgements

This research has been conducted using the UK Biobank Resource under Application Number 44584.

## Abbreviations

T2D: Type 2 Diabetes
BMI: Body mass index
WHR: waist-to-hip ratio
AST:ALT: ratio of aspartate aminotransferase to alanine aminotransferase
FIB-4: Fibrosis-4 index
Liver PDFF: Liver percentage density fat fraction
MUR: Mass univariate regression
TFCE: Threshold-free cluster enhancement
SPMs: Statistical parametric maps
S2S: Surface-to-surface.

## Notes

### Competing Interest Statement

MC and EPS are employees of Calico Life Sciences LLC

### Funding Statement

This research was funded by Calico Life Sciences LLC.

### Author Declarations

Participant data from the UK Biobank cohort was obtained through UK Biobank Access Application number 44584. The UK Biobank has approval from the North West Multi-Centre Research Ethics Committee (REC reference: 11/NW/0382). All methods were performed in accordance with the relevant guidelines and regulations, and informed consent was obtained from all participants. Researchers may apply to use the UKBB data resource by submitting a health-related research proposal (https://www.ukbiobank.ac.uk).

### Summary of Updates

A futher predictive analysis was conducted and Table 4 was added.

