## Supplementary_Material for "Liver Shape Analysis using Statistical Parametric Maps at Population Scale"

### Disease Categories

Disease categories of interest, including liver disease and type-2 diabetes (T2D), were selected based on their influence in body composition and organ health, as well as their frequency within the UK Biobank. They were defined based on recorded hospital episode statistics (HES) data and self-reported information at the time and before the baseline imaging visit. Specifically, the (International Classification of Disease) ICD codes for liver disease were taken from [1] and the self-reported codes used were "liver failure/cirrhosis", "infective/viral hepatitis", "alcoholic liver disease/alcoholic cirrhosis" and "liver/biliary/pancreas problem" and the codes for T2D were selected based on the ICD codes for "non-insulin-dependent diabetes mellitus" and the self-reported code for "diabetes" and "type-2 diabetes" [2]. Liver disease and type-2 diabetes diagnosed or reported after the baseline imaging visit was defined based on the occurrence of the 3-character ICD10 codes (see [2]) mapped from the various sources, the date of first diagnosis made available as "first occurrence" data (Category 1712) and based on self-reported codes after the baseline imaging visit.

Supplementary Tables

| | 20-participant template | | 100-participant template | | 200-participant template | |
|---|---|---|---|---|---|---|
| | **Male** | **Female** | **Male** | **Female** | **Male** | **Female** |
| **Age** ($yrs.$) | 52.7 ± 1.89 (50, 57) | 51.4 ± 2.46 (47, 55) | 53.38 ± 3.06 (48, 62) | 50.72 ± 2.37 (46, 59) | 55.13 ± 3.56 (48, 62) | 52.39 ± 3.35 (46, 62) |
| **Weight** ($kg$) | 68.95 ± 5.61 (61.4, 78.7) | 55.14 ± 5.2 (46.2, 63.2) | 70.65 ± 6.43 (57.5, 90.1) | 58.96 ± 5.85 (46.2, 72.4) | 70.92 ± 6.63 (57.5, 90.1) | 60.30 ± 6.16 (46.2, 76.8) |
| **Height** ($m$) | 1.75 ± 0.05 (1.67, 1.84) | 1.64 ± 0.07 (1.53, 1.74) | 1.61 ± 0.07 (1.77, 1.92) | 1.65 ± 0.06 (1.51, 1.78) | 1.78 ± 0.07 (1.61, 1.93) | 1.66 ± 0.06 (1.51, 1.80) |
| **BMI** ($kg/m^2$) | 22.51 ± 1.47 (20.1, 24) | 20.5 ± 1.6 (19.2, 24.8) | 22.53 ± 1.58 (18.5, 24.9) | 21.71 ± 1.47 (18.6, 24.8) | 22.41 ± 1.50 (18.5, 24.9) | 21.94 ± 1.6 (18.6, 24.8) |
| **Waist circum-ference** ($cm$) | 82.7 ± 5.83 (71, 92) | 71.5 ± 7.58 (63, 91) | 81.73 ± 5.89 (68, 92) | 71.7 ± 5.96 (61, 91) | 81.79 ± 5.78 (68, 93) | 71.82 ± 5.86 (60, 91) |
| **Hip circumference** ($cm$) | 93.1 ± 4.01 (86, 98) | 91 ± 3.23 (86, 98) | 93.88 ± 4.42 (86, 103) | 92.32 ± 4.29 (83, 100) | 94.23 ± 4.73 (85, 111) | 93.05 ± 4.68 (82, 103) |
| **WHR** | 0.89 ± 0.05 (0.72, 0.95) | 0.78 ± 0.06 (0.72, 0.93) | 0.87 ± 0.05 (0.74, 0.98) | 0.78 ± 0.05 (0.68, 0.93) | 0.87 ± 0.05 (0.74, 0.98) | 0.77 ± 0.05 (0.67, 0.93) |
| **AST:ALT** | 1.03 ± 0.25 (0.61, 1.34) | 1.37 ± 0.26 (0.99, 1.95) | 1.3 ± 0.33 (0.61, 1.95) | 1.38 ± 0.27 (0.83, 2.30) | 1.36 ± 0.39 (0.62, 2.81) | 1.47 ± 0.39 (0.83, 3.58) |
| **FIB-4** | 1.14 ± 0.35 (0.70, 1.70) | 1.00 ± 0.27 (0.59, 1.35) | 1.26 ± 0.38 (0.59, 2.46) | 0.95 ± 0.28 (0.52, 1.97) | 1.26 ± 0.42 (0.59, 2.59) | 1.01 ± 0.32 (0.45, 1.97) |
| **Liver PDFF** (%) | 2.05 ± 0.40 (1.60, 2.90) | 1.96 ± 0.46 (1.21, 2.58) | 2.00 ± 0.33 (1.21, 2.90) | 1.73 ± 0.36 (1.14, 2.58) | 2.09 ± 0.39 (1.21, 2.99) | 1.80 ± 0.40 (1.14, 2.93) |
| **Liver Iron** ($mg/g$) | 1.19 ± 0.16 (1.05, 1.49) | 1.06 ± 0.11 (0.91, 1.30) | 1.16 ± 0.14 (0.95, 1.52) | 1.07 ± 0.16 (0.85, 1.59) | 1.20 ± 0.20 (0.86, 1.84) | 1.06 ± 0.13 (0.85, 1.59) |

| | 20-participant template | | 100-participant template | | 200-participant template | |
|---|---|---|---|---|---|---|
| | **Male** | **Female** | **Male** | **Female** | **Male** | **Female** |
| **Age** ($yrs.$) | 52.7 ± 1.89 (50, 57) | 51.4 ± 2.46 (47, 55) | 53.38 ± 3.06 (48, 62) | 50.72 ± 2.37 (46, 59) | 55.13 ± 3.56 (48, 62) | 52.39 ± 3.35 (46, 62) |
| **Liver Volume** ($l$) | 1.21 ± 0.17 (0.97, 1.61) | 1.21 ± 0.16 (0.96, 1.45) | 1.39 ± 0.18 (0.97, 1.69) | 1.21 ± 0.13 (0.96, 1.47) | 1.41 ± 0.20 (0.97, 1.96) | 1.24 ± 0.14 (0.96, 1.59) |

**Table S1.** Summary statistics (mean ± standard deviation, minimum and maximum values) of a gender-balanced cohort for the template construction with 20, 100 and 200 participants. BMI: body mass index, WHR: waist-to-hip ratio, AST:ALT: aspartate aminotransferase/alanine aminotransferase ratio, FIB-4: Fibrosis-4 score, Liver PDFF: Liver percentage density fat fraction, T2D: type-2 diabetes.

| | **Full Cohort** | **Male** (N=195) | **Female** (N=305) |
|---|---|---|---|
| **Age** ($yrs.$) | 55.42 ± 3.86 (46,62) | 55.5 ± 3.89 (47,62) | 55.36 ± 3.84 (46,62) |
| **Weight** ($kg$) | 64.99 ± 8.33 (46.20, 90.10) | 71.67 ± 6.85 (52.80,90.10) | 60.72 ± 6.08 (46.20, 79.40) |
| **Height** ($m$) | 1.71 ± 0.09 (1.50, 1.97) | 1.83 ± 0.07 (1.59, 1.97) | 1.66 ± 0.06 (1.50, 1.81) |
| **BMI** ($kg/m^2$) | 22.26 ± 1.59 (18.51, 24.90) | 22.53 ± 1.45 (18.51, 24.90) | 22.1 ± 1.66 (18.51, 24.83) |
| **Waist circumference** ($cm$) | 76.45 ± 7.45 (59, 93) | 82.13 ± 5.41 (67, 93) | 72.82 ± 6.21 (59, 93) |

| | | | |
|---|---|---|---|
| **Hip circumference** $(cm)$ | 94.24 ± 4.97 (81, 111) | 94.28 ± 4.48 (82, 111) | 94.21 ± 5.27 (81, 110) |
| **WHR** | 0.81 ± 0.07 (0.63, 0.99) | 0.87 ± 0.05 (0.74, 0.99) | 0.77 ± 0.05 (0.63, 0.93) |
| **AST:ALT** | 1.47 ± 0.39 (0.58, 3.33) | 1.36 ± 0.35 (0.61, 2.81) | 1.53 ± 0.39 (0.58, 3.33) |
| **FIB-4** | 1.18 ± 0.46 (0.48, 5.30) | 1.25 ± 0.40 (0.55, 2.59) | 1.14 ± 0.49 (0.48, 5.30) |
| **Liver PDFF** (%) | 2.09 ± 0.42 (1.15, 2.99) | 2.22 ± 0.40 (1.21, 2.99) | 2.02 ± 0.41 (1.15, 2.97) |
| **Liver Iron** $(mg/g)$ | 1.16 ± 0.20 (0.86, 2.55) | 1.21 ± 0.23 (0.86, 2.55) | 1.13 ± 0.17 (0.87, 1.87) |
| **Liver Volume** $(l)$ | 1.30 ± 0.21 (0.89, 2.27) | 1.41 ± 0.22 (0.97, 2.27) | 1.23 ± 0.16 (0.89, 1.66) |

**Table S2.** Summary statistics (mean, standard deviation, minimum and maximum values) of a 500-participants cohort. BMI: body mass index, WHR: waist-to-hip ratio, AST:ALT: aspartate aminotransferase/alanine aminotransferase ratio, FIB-4: Fibrosis-4 score, Liver PDFF: Liver percentage density fat fraction, T2D: type-2 diabetes.

| | **Full Cohort** | **Male** (N=16,264) | **Female** (N=17,170) |
|---|---|---|---|
| **Caucasian** $(N)$ | 32,623 | 15,829 | 16,794 |
| **Age** $(yrs.)$ | 64.26 ± 7.73 | 64.98 ± 7.82 | 63.57 ± 7.57 |

| | | | |
|---|---|---|---|
| | (44, 82) | (44, 81) | (45, 82) |
| **Weight** $(kg)$ | 76.14 ± 15.04<br>(34.30, 169.70) | 83.68 ± 13.19<br>(44.50,169.20) | 69 ± 13.07<br>(34.30,169.70) |
| **Height** $(m)$ | 1.69 ± 0.09<br>(1.34, 2.04) | 1.76 ± 0.07<br>(1.50, 2.04) | 1.63 ± 0.06<br>(1.34, 1.95) |
| **BMI** $(kg/m^2)$ | 26.49 ± 4.35<br>(14.08, 55.10) | 26.96 ± 3.85<br>(16.48, 53.91) | 26.05 ± 4.73<br>(14.08, 55.10) |
| **Waist circumference** $(cm)$ | 88.43 ± 12.65<br>(53, 184) | 94.36 ± 10.65<br>(63, 184) | 82.81 ± 11.81<br>(53, 144) |
| **Hip circumference** $(cm)$ | 100.79 ± 8.68<br>(71, 157) | 100.71 ± 7.26<br>(76.2, 152) | 100.86 ± 9.83<br>(71, 157) |
| **WHR** | 0.88 ± 0.09<br>(0.61, 1.47) | 0.94 ± 0.06<br>(0.64, 1.47) | 0.82 ± 0.07<br>(0.61, 1.22) |
| **AST:ALT** | 1.27 ± 0.42<br>(0.16, 6.33) | 1.15 ± 0.39<br>(0.30, 6.33) | 1.37 ± 0.43<br>(0.16, 5.99) |
| **FIB-4** | 1.31 ± 0.59<br>(0.17, 39.63) | 1.39 ± 0.6<br>(0.32, 29.49) | 1.23 ± 0.57<br>(0.17, 39.63) |
| **Liver PDFF** (%) | 4.95 ± 4.91<br>(0.53, 50.30) | 5.58 ± 5.1<br>(0.53, 50.30) | 4.35 ± 4.65<br>(0.83, 43.97) |
| **Liver Iron** $(mg/g)$ | 1.21 ± 0.28<br>(0.22, 7.29) | 1.22 ± 0.30<br>(0.34, 6.99) | 1.20 ± 0.26<br>(0.22, 7.29) |
| **Liver Volume** $(l)$ | 1.41 ± 0.31 | 1.53 ± 0.31 | 1.31 ± 0.27 |

| | | | |
|---|---|---|---|
| | (0.64, 4.18) | (0.71, 4.08) | (0.64, 4.18) |

**Table S3.** Summary statistics (mean ± standard deviation, minimum and maximum values) for continuous variables and counts for discrete variables in the full cohort (N=33,434). BMI: body mass index, WHR: waist-to-hip ratio, AST:ALT: aspartate aminotransferase/alanine aminotransferase ratio, FIB-4: Fibrosis-4 score, Liver PDFF: Liver percentage density fat fraction, T2D: type-2 diabetes.

| | **Liver Disease** | **T2D** |
|---|---|---|
| **Sex** | 207F / 242M | 599F / 1,181M |
| **Caucasian** $(N)$ | 435 | 1,669 |
| **Age** $(yrs.)$ | 65.75 ± 7.47<br>(48, 81) | 66.63 ± 7.34<br>(46, 82) |
| **Weight** $(kg)$ | 81.32 ± 15.53<br>(46.40, 140.40) | 86.46 ± 16.81<br>(41.90, 169.20) |
| **Height** $(m)$ | 1.70 ± 0.09<br>(1.50, 1.95) | 1.71 ± 0.09<br>(1.44, 1.97) |
| **BMI** $(kg/m^2)$ | 28.25 ± 4.66<br>(18.59, 49.16) | 29.62 ± 5.14<br>(18.31, 54.13) |
| **Waist circumference** $(cm)$ | 94.04 ± 12.83<br>(58, 135) | 98.92 ± 13.15<br>(59, 152) |
| **Hip circumference** $(cm)$ | 103.78 ± 9.21<br>(77, 152) | 104.84 ± 10.34<br>(76, 155) |

| | | |
|---|---|---|
| **WHR** | 0.91 ± 0.09<br>(0.66. 1.13) | 0.94 ± 0.09<br>(0.66, 1.26) |
| **AST:ALT** | 1.16 ± 0.42<br>(0.41, 2.76) | 1.03 ± 0.37<br>(0.40, 5.71) |
| **FIB-4** | 1.47 ± 0.76<br>(0.39, 7.92) | 1.33 ± 0.58<br>(0.43, 8.50) |
| **Liver PDFF** (%) | 7.04 ± 6.87<br>(0.92, 42.16) | 8.94 ± 7.23<br>(0.88, 40.36) |
| **Liver Iron** ($mg/g$) | 1.21 ± 0.34<br>(0.55, 5.32) | 1.16 ± 0.22<br>(0.34, 5.22) |
| **Liver Volume** ($l$) | 1.51 ± 0.39<br>(0.64, 3.39) | 1.70 ± 0.46<br>(0.78, 4.18) |

**Table S4.** Summary statistics (mean ± standard deviation, minimum and maximum values) for continuous variables and counts for discrete variables in the liver disease (N=449) and in the T2D (N=1,780) cohort, diagnosed at baseline imaging visit. F: Female; M: Male; BMI: body mass index, WHR: waist-to-hip ratio, AST:ALT: aspartate aminotransferase/alanine aminotransferase ratio, FIB-4: Fibrosis-4 score, Liver PDFF: Liver percentage density fat fraction, T2D: type-2 diabetes.

| | Sample 1 | Sample 2 | Sample 3 | Sample 4 | Sample 5 |
|---|---|---|---|---|---|
| Sample 1 | | | | | |
| Sample 2 | 0.924 | | | | |
| Sample 3 | 0.942 | 0.947 | | | |

| | | | | |
|---|---|---|---|---|
| Sample 4 | 0.933 | 0.940 | 0.951 | |
| Sample 5 | 0.917 | 0.939 | 0.941 | 0.938 |

**Table S5.** Dice similarity scores on the template images from 20 participants, repeated 5 times with different samples from the population (Sample 1 - Sample 5).

| | $\hat{\beta} < 0$ | | $\hat{\beta} > 0$ | | Total |
|---|---|---|---|---|---|
| | Beta coefficients | Significance area | Beta coefficients | Significance area | Significance area |
| **Age** $(yrs.)$ | -0.11 (0.06) | 96.58% | 0.02 (0.04) | 1.56% | 98.13% |
| **BMI** $(kg/m^2)$ | -0.08 (0.07) | 1.60% | 0.30 (0.22) | 97.84% | 99.44% |
| **WHR** | -3.87 (4.02) | 34.02% | 3.87 (3.64) | 58.10% | 92.12% |
| **AST:ALT** | -0.32 (0.32) | 35.33% | 0.30 (0.29) | 48.01% | 83.34% |
| **FIB-4** | -0.22 (0.17) | 82.48% | 0.23 (0.13) | 2.14% | 84.62% |
| **Liver PDFF** (%) | -0.03 (0.01) | 0.16% | 0.26 (0.10) | 99.64% | 99.81% |
| **Liver Iron** $(mg/g)$ | -0.62 (0.73) | 55.59% | 0.37 (0.32) | 23.15% | 78.74% |
| **Liver disease** | -2.40 (2.90) | 14.33% | 2.30 (2.31) | 14.01% | 28.34% |
| **T2D** | -0.59 (0.77) | 4.93% | 2.43 (1.94) | 85.85% | 90.78% |
| **Age * Liver disease** | ns | ns | ns | ns | ns |

| | | | | | |
|---|---|---|---|---|---|
| **Liver PDFF * Liver disease** | ns | ns | ns | ns | ns |
| **Age * T2D** | -0.04 (0.02) | 64.48% | 0 | 0% | 64.48% |
| **Liver PDFF * T2D** | -0.06 (0.04) | 6.27% | 0.10 (0.08) | 82.33% | 88.60% |

**Table S6.** Significance areas for covariates in the MUR model between the anthropometric covariates and liver IDPs (N=33,434) in model 3. The total area has been split into areas of positive and negative associations. The regression coefficients are presented as median (interquartile range - IQR) and the significance areas as a percentage (%) of the vertices. Where BMI: body mass index, WHR: waist-to-hip ratio, AST:ALT: aspartate aminotransferase/alanine aminotransferase ratio, FIB-4: Fibrosis-4 score, Liver PDFF: Liver percentage density fat fraction, T2D: type-2 diabetes.

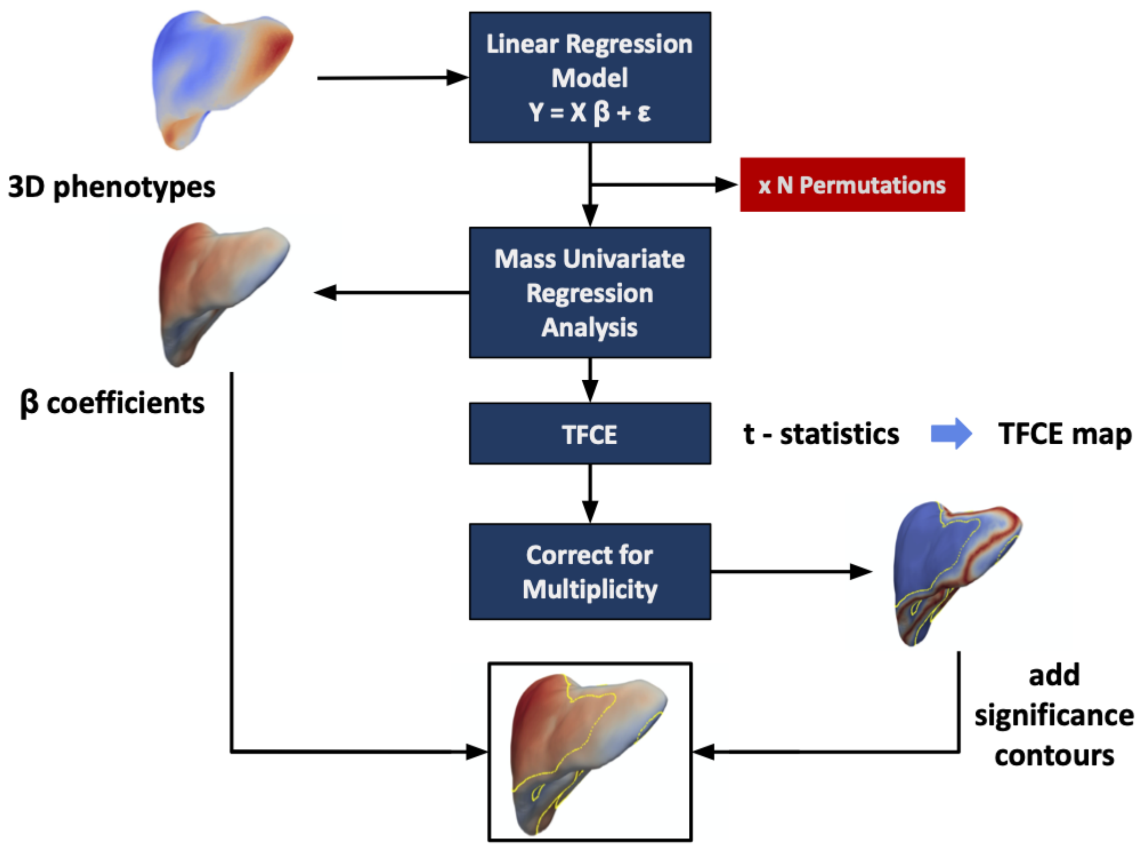

**Figure S1.** Flow diagram for the mass univariate regression (MUR) analysis of three-dimensional phenotypes. The phenotypes are used to construct the linear regression model. MUR analysis produces parameter estimates ($\hat{\beta}$) and their null distribution via permutation. Threshold free cluster enhancement (TFCE) is applied to the $t$-statistics from the regression analysis to produce a significance threshold. The associated $p$-values are corrected for multiple comparisons and mapped onto the mesh for visualisation. This diagram was modified from [3].

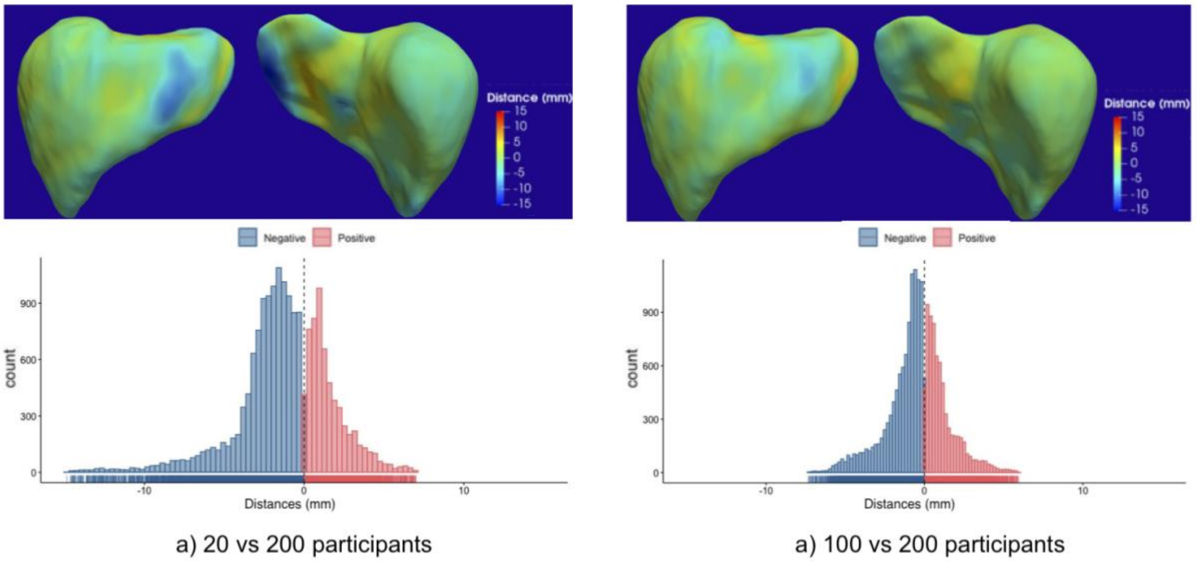

**Figure S2.** Three-dimensional maps and histograms showing the differences (in mm) between each template constructed using a sub-population of 20 vs. 200 (a) and 100 vs 200 participants (b). Areas with positive distances indicate that the observed template is bigger than the 200-participant template and negative distances indicate that the observed template is smaller than the 200-participant template.

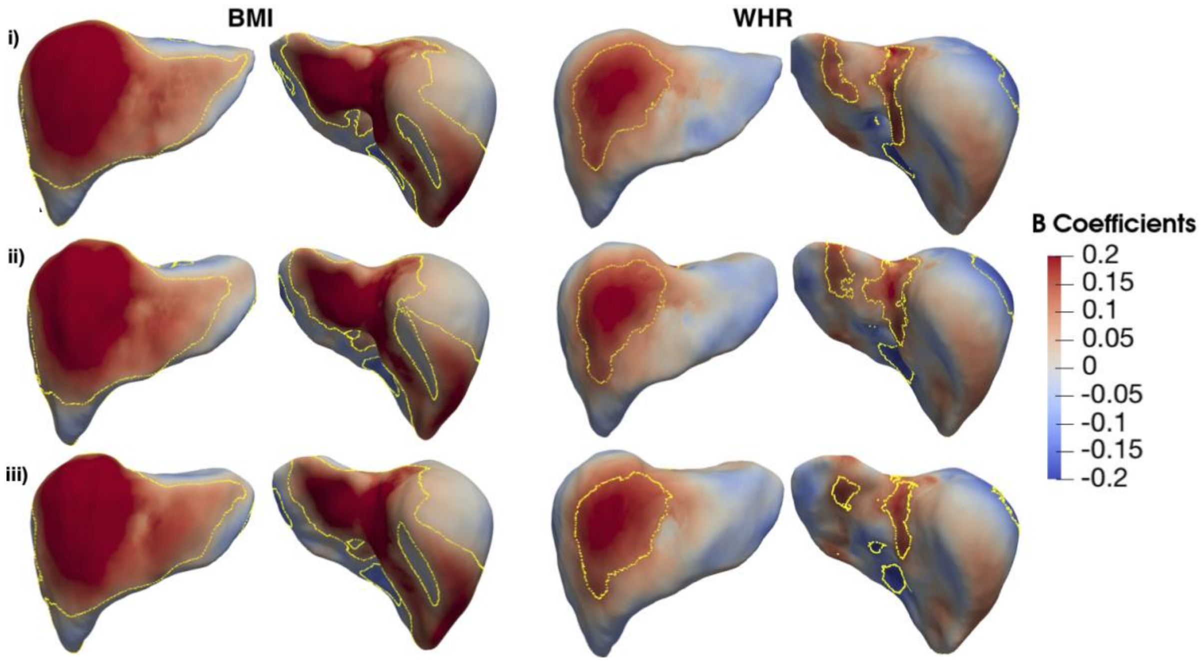

**Figure S3.** Three-dimensional statistical parametric maps of liver morphology, projections are shown for each SPM providing anterior (left) and posterior (right) views of the liver. The SPMs show the local strength of association between (i) BMI and WHR with S2S values on a 500-participants cohort where the templates constructed from 20 (i), 100 (ii) and 200 (iii) participants were registered to each participant surface meshes. Yellow contour lines indicate statistically significant regions after correction for multiple testing ($p < 0.05$), with positive associations in red and negative associations in blue. Standardised regression coefficients are shown with units in standard deviations for each covariate.

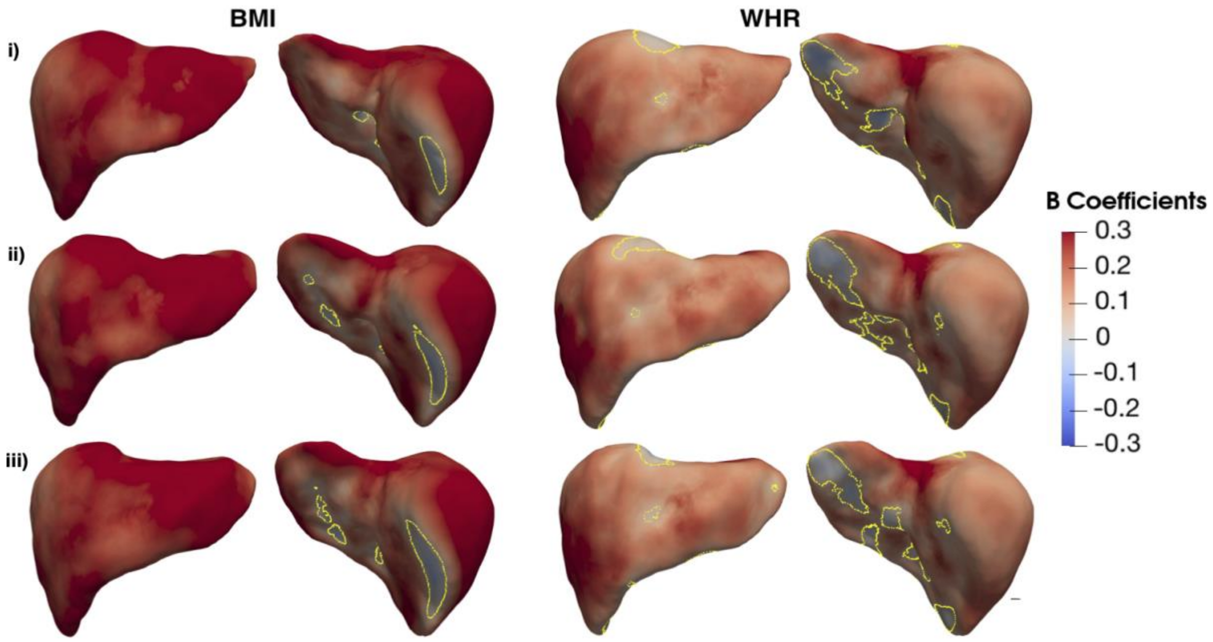

**Figure S4.** Three-dimensional statistical parametric maps of liver morphology, projections are shown for each SPM providing anterior (left) and posterior (right) views of the liver. The SPMs show the local strength of association between BMI and WHR with S2S values on a cohort with liver disease (N=449), where the templates constructed from 20 (i), 100 (ii) and 200 (iii) participants were registered to each participant surface meshes. Yellow contour lines indicate statistically significant regions ($p < 0.05$) after correction for multiple testing, with positive associations in red and negative associations in blue. Standardised regression coefficients are shown with units in standard deviations for each covariate.

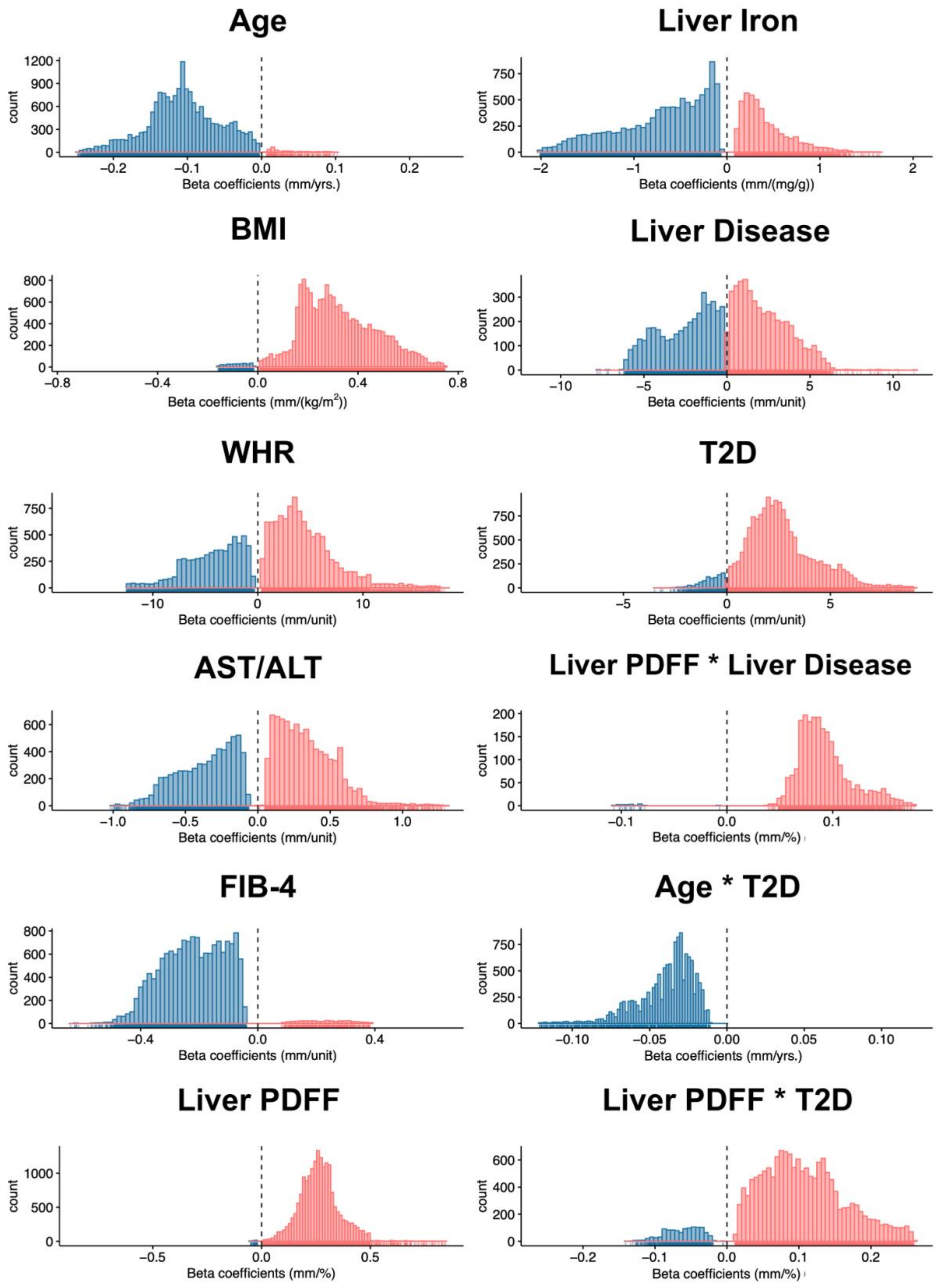
Age
count
Beta coefficients (mm/yrs.)
Liver Iron
count
Beta coefficients (mm/(mg/g))
BMI
count
Beta coefficients (mm/(kg/m$^2$))
Liver Disease
count
Beta coefficients (mm/unit)
WHR
count
Beta coefficients (mm/unit)
T2D
count
Beta coefficients (mm/unit)
AST/ALT
count
Beta coefficients (mm/unit)
Liver PDFF * Liver Disease
count
Beta coefficients (mm/%)
FIB-4
count
Beta coefficients (mm/unit)
Age * T2D
count
Beta coefficients (mm/yrs.)
Liver PDFF
count
Beta coefficients (mm/%)
Liver PDFF * T2D
count
Beta coefficients (mm/%)

**Figure S5.** Histograms showing the statistically significant regression coefficients across the vertices (~18,000) for each covariate in model 2 on the full cohort (N=33,434) with positive associations in red and negative associations in blue. Regression coefficients are provided in their physical units in order to facilitate interpretation.

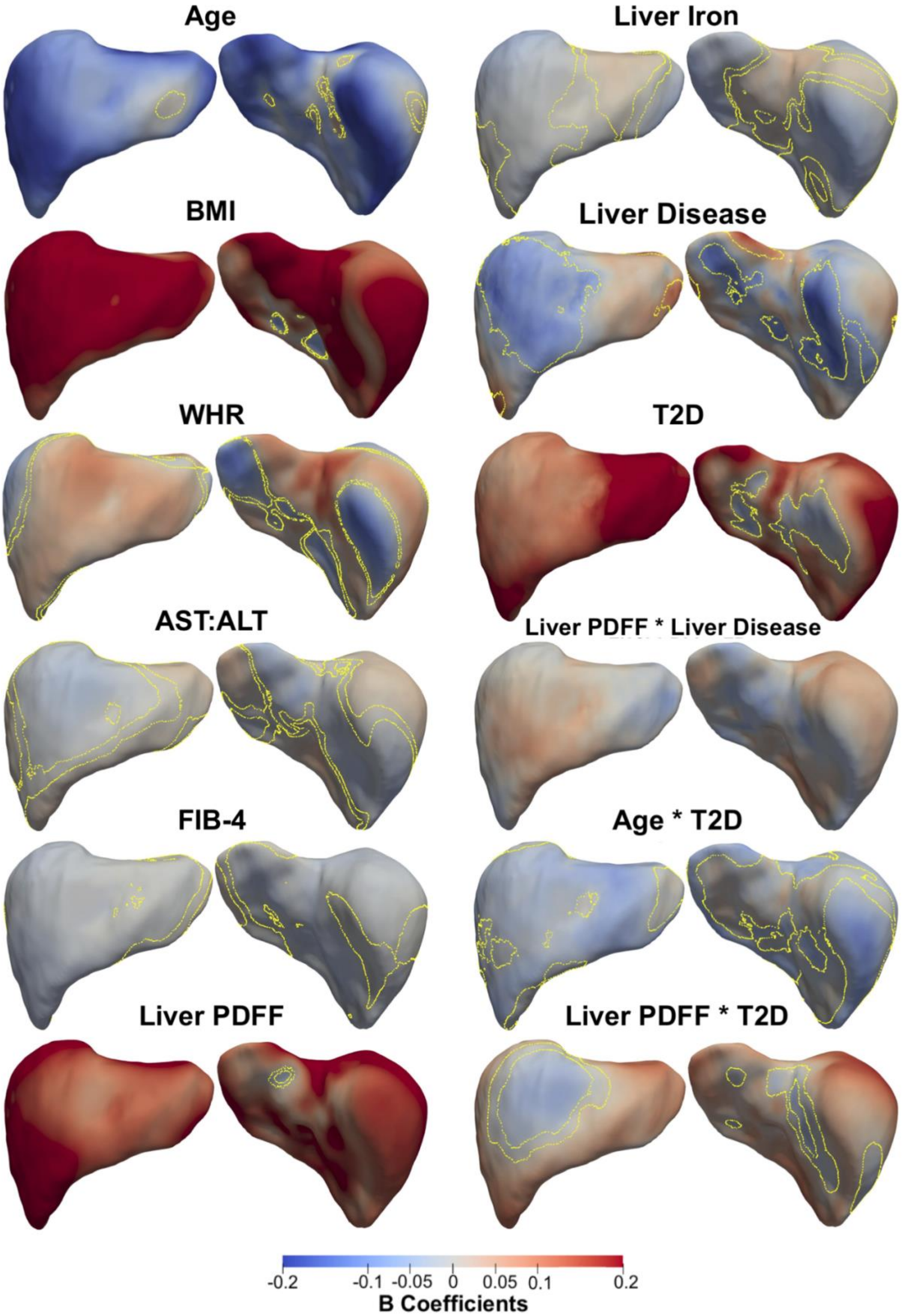

Age
Liver Iron
BMI
Liver Disease
WHR
T2D
AST:ALT
Liver PDFF * Liver Disease
FIB-4
Age * T2D
Liver PDFF
Liver PDFF * T2D
-0.2
-0.1
-0.05
0
0.05
0.1
0.2
B Coefficients

**Figure S6.** Three-dimensional statistical parametric maps (SPMs) of liver morphology, two projections are shown for each SPM providing anterior (left) and posterior (right) views of the liver. The SPMs show the local strength of association for each covariate in model 3 with S2S distances on the full cohort (N=33,434). Yellow contour lines indicate the boundary between statistically significant regions ($p < 0.05$) after correction for multiple testing, with positive associations in red and negative associations in blue. Standardised regression coefficients are shown with units in standard deviations for each covariate. Where BMI: body mass index, WHR: waist-to-hip ratio, AST:ALT: aspartate aminotransferase/alanine aminotransferase ratio, FIB-4: Fibrosis-4 score, Liver PDFF: Liver percentage density fat fraction, T2D: type-2 diabetes.

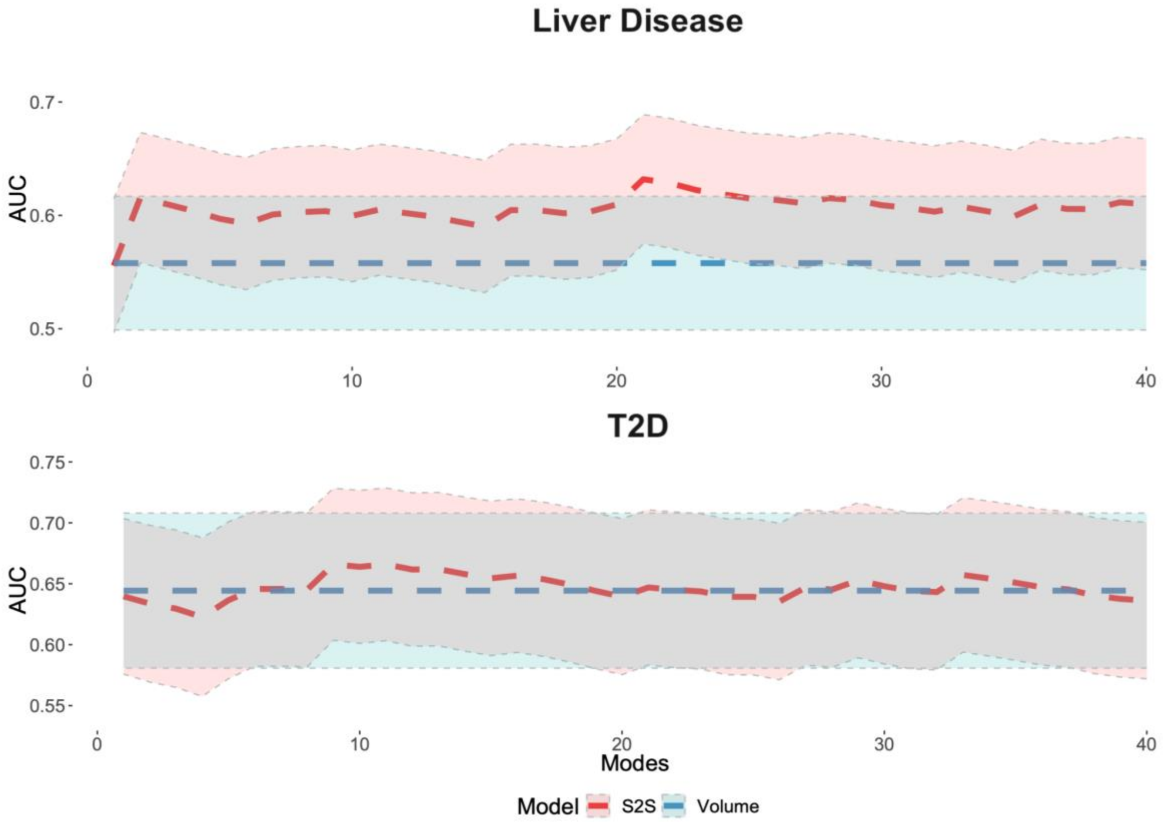

**Figure S7.** Cumulative area under the curve (AUC) and 95% confidence intervals (CI) for the predictive model using liver volume and the predictive model using S2S distances with increasing numbers of modes from 1 to 40, for both liver disease (top plot, N=364) and T2D (bottom plot, N=288) case-control groups.
